# Investigation of Compatibility of SARS-CoV-2 RT-PCR Kits Containing Different Gene Targets During COVID-19 Pandemic

**DOI:** 10.1101/2020.06.17.20133967

**Authors:** Figen Sarıgul, Osman Doluca, Sıla Akhan, Murat Sayan

## Abstract

**Aim:** In the diagnosis of severe acute respiratory syndrome coronavirus 2 (SARS-CoV-2), reverse transcriptase-polymerase chain reaction (RT-PCR) technique is often used. We evaluated the compatibility of SARS-CoV-2 RT-PCR kits containing different gene targets during the pandemic.

**Materials & methods:** **S**amples were tested by Bio-Speddy® (RdRp gene) and Diagnovital® (RdRp+E genes). The correlation between two assays were determined by Deming regression and chi-square heatmap analyses.

**Results:** Diagnovital PCR kit showed in a constricted range and conveniently exponential amplification curves than Bio-Speedy PCR kit. While the correlation increased when a secondary biomarker was added to the kit.

**Conclusion:** In an unknown sample, using together different PCR kits that target different genes during the pandemic situation may provide a more accurate diagnosis of SARS-CoV-2.

## INTRODUCTION

The disease-causing factor was determined to be a new coronavirus after reports of pneumonia cases identified in Wuhan Province of China in December 2019 to the World Health Organization (WHO) regional office. It was declared as a pandemic by WHO on March 11, 2020, after the cases spread across the world. The new virus was an etiological agent of pandemic coronavirus disease 2019 (COVID-19), and since the genome of the new virus was the same as severe acute respiratory syndrome coronavirus (SARS-CoV), it was named as SARS-CoV-2 (1). SARS-CoV-2 was found to have a much higher transmission rate than known human coronavirus strains, while also causing damage to the lung tissue, causing respiratory failure and potentially leading to death. Individuals over 65 years of age, smokers, and people with chronic diseases such as hypertension, diabetes, kidney failure were more severely affected. Patients often showed signs of dry cough, high fever, and shortness of breath, while some patients with abdominal pain, diarrhea, and headache have also been reported. It was understood that some infected individuals remained asymptomatic. As of the end of May 2020, the number of cases reported worldwide as SARS-CoV-2 positive has exceeded 4.5 million, and deaths exceeded 300,000 (2,3). In Turkey, the first cases in which the WHO declared a pandemic, March 11, 2020 were seen, and 157.814 cases and 4369 death were present at the end of May 2020 (4). According to new phylogenetic analysis of the virus in Turkey; the introduction of the virus into the country for the first time is earlier than the first reported case of infection, and the virus was found to have many independent international entries into the country (5)

First of all, to prevent COVID-19 pandemic, early detection and isolation of asymptomatic and symptomatic infections are necessary. Diagnostic tests information for SARS-CoV-2 is still developing, and a clear understanding of the nature of the tests and interpretation of their findings is essential. During the COVID-19 pandemic, in the diagnosis of SARS-CoV-2, reverse transcriptase-polymerase chain reaction (RT-PCR) and IgM and IgG enzyme-linked immunosorbent assay tests are often used (6,7). Additionally, it remains unclear whether RT-PCR is the gold standard and whether false positive or false negative results are common. Centers for Disease Control and Prevention (CDC) recommends the collection of oropharyngeal and nasopharyngeal swab specimens to test for SARS-CoV-2. (8). Various RNA gene targets are used by different manufacturers; most tests target envelopes (E), spike (S), nucleocapsid (N), RNA-linked RNA polymerase (RdRp), and ORF1 genes 1 or more (9). Among these assays, the RdRp assay had the highest analytical sensitivity (3.8 RNA copies/ml reaction at 95% detection probability) (10). In Turkey, the diagnosis was made according to the recommendations given by “COVID-19 Diagnosis and Treatment Guide” published by the Turkish Ministry of Health (11). RT-PCR diagnostic kit for detecting SARS-CoV-2 in respiratory tract samples has been developed by the Ministry of Health General Directorate of Public Health Microbiology Reference Laboratories and Biological Products Department Virology Laboratory. From the beginning of the COVID-19 epidemic in Turkey, RdRp assay developed in-house and applied in all regions of the country.

In COVID-19 infection, viral RNA in the nasopharyngeal swab, measured by the threshold cycle (Ct), can be detected in most individuals a week before the onset of symptoms and peaked in the first week of symptom onset. Ct is the number of replication cycles required to produce a fluorescent signal with low Ct values indicating high viral RNA loads. Those with PCR positive will have a Ct value of less than 40. PCR positivity begins to decrease until the 3rd week and then becomes undetectable (12). Failure of the sampling process to coincide with the appropriate time and maloperation of nasopharyngeal swabs can cause false negative results. Most RT-PCR tests have a specificity of 100% because their primary design is specific to the genome sequence of SARS-CoV-2. False positive results may occasionally occur due to technical errors and reagent contamination (13,14).

RdRp assays had been implemented in 30 laboratories in Europe (15). However, this method requires improvement as a molecular diagnostic test used by clinical laboratories. The WHO recommended the detection of at least two different targets on the SARS-CoV-2 genome (16). It is also recommended that E gene assay as a primary screening tool followed by the RdRp gene assay as confirmatory tests (17,18). However, there are problems in the ongoing diagnostic processes with PCR kits. We do not have sufficient information on the diagnostic powers, validations, and standards of the kits used. Questions such as comparative analysis of PCR diagnostic kits, analysis compatibility with each other, and which genes of SARS-CoV-2 cause in diagnostic processes await answers. In the COVID-19 pandemic, we aimed that we would understand the problems encountered during the diagnosis process with PCR kits and obtain data on how to improve the diagnostic process.

International external quality controls of the kits used have not been established yet. Therefore, an unknown sample is correlated with the close distances of the compared kits when measuring the correct diagnosis. Also, PCR tests used in pandemics are qualitative for an early and accurate diagnosis. Here we evaluated the compatibility of two different multiplex RT-PCR methodologies used in the simultaneous detection of two regions of the SARS-CoV-2 genome (E and RdRp genes) and the RdRp gene.

## METHODOLOGY

A total of 96 patients oro/nasopharyngeal swab samples from different hospitals in Kocaeli city (Kocaeli State Hospital, Seka State Hospital, Gebze Darıca State Hospital and Gölcük State Hospital) which were sending for SARS-CoV-2 diagnosis to PCR unit of Kocaeli University were included to this study between March and April 2020. All samples were studied by a comparison of two different RT-PCR kits produced in Turkey.

One of the RT-PCR kits is the Bio-Speddy® (Bioeksen R&D Technologies Inc. COVID-19 RT-qPCR Detection Kit v2.0, Istanbul-Turkey) determined valuable by the “Turkish Ministry of Health” and used throughout COVID-19 pandemic. The other kit; Diagnovital® (RTA Laboratories Inc, SARS-CoV-2 Real-Time PCR Kit v2.0 Istanbul-Turkey) is a commercial kit and mainly exported to European and Middle East-North Africa zones during the pandemic. Each RT-PCR kit production followed CDC’s and WHO’s detection guidelines (19,20). However, each kits are included in the WHO Emergency Use Listing for SARS-CoV-2 in vitro diagnostic products (https://www.who.int/diagnostics_laboratory/EUL/en/). Viral RNA extraction from samples were performed according to the manufacturer’s instructions. For automated viral nucleic acid extraction processing Qiagen - EZ 1 Advanced XL platform (Qiagen, Hilden Germany) and for PCR assay run Qiagen - Rotorgene Q thermal cycler platform (Qiagen, Hilden Germany) was used. Limit of detection values of the kits of analytical sensitivity belonging in 95% confidence. A negative (human specimen control) was included in every RNA extraction procedure, and a non-template (water) control was included in every RT-PCR run. An internal control amplification was performed to monitor RNA extraction and RT-PCR quality. Characteristics of the SARS-CoV-2 RT-PCR kits are shown in Table 1.

**Table 1.**
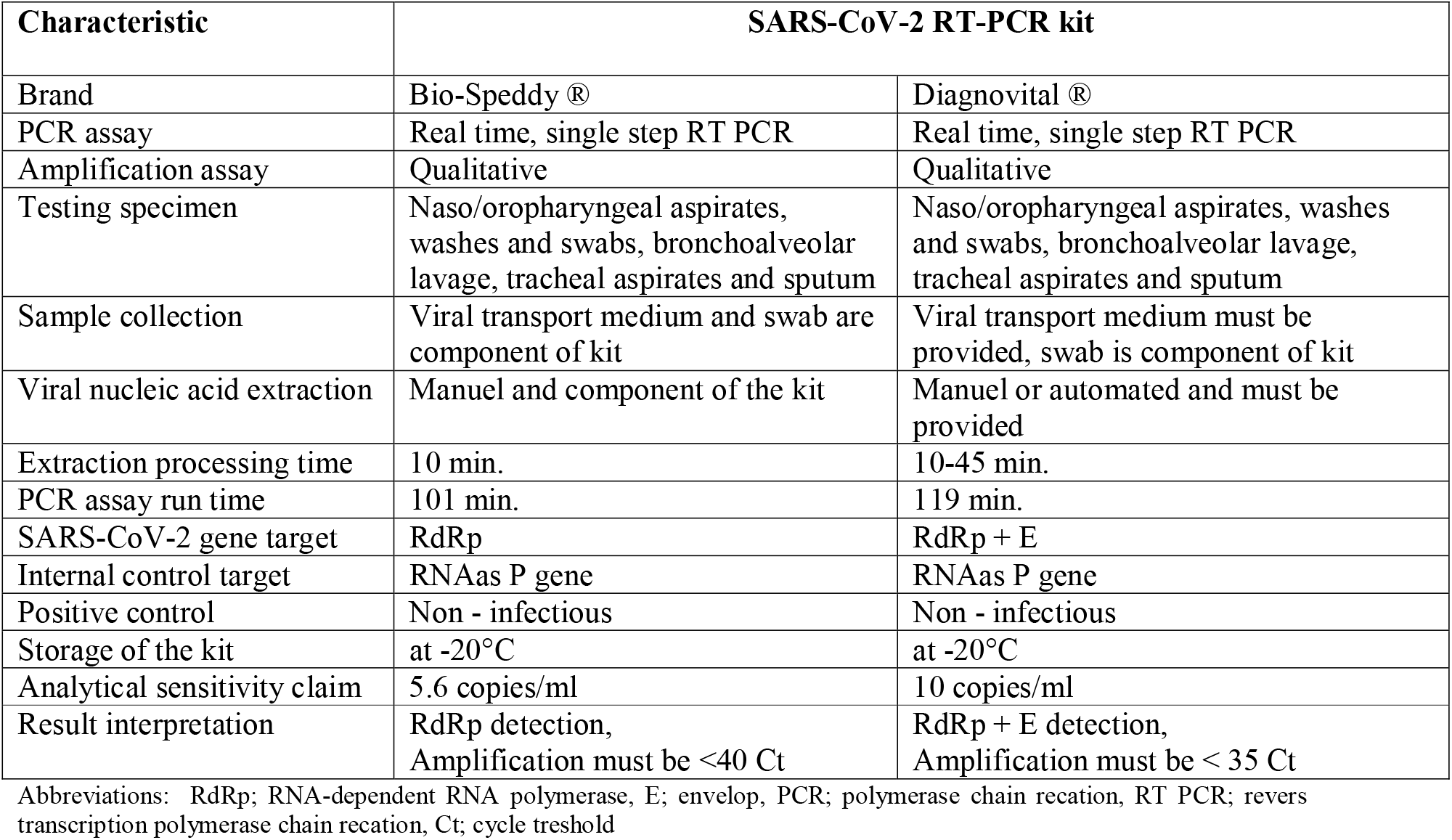
Comparison of characteristics the SARS-CoV-2 PCR kits which analysed in the study

Swab materials available for sample collection for COVID-19 based on the CDC Interim Guidelines for Collecting, Handling, and Testing Clinical Specimens from Patients Under Investigation for COVID-19 (21). Oropharyngeal/nasopharyngeal samples were collected from the patients by synthetic fiber swabs with plastic shafts (Citotest Scientific Co, Haimen City, P.R.China). The swabs were placed in 3 mL sterile viral transport media (Citotest Scientific Co, Haimen City, P.R.China) during the collection and transferred with biohazard specimen bag. After the samples were taken, they were transferred to the center and tested shortly within the 2 hours. Samples were vortexed for 3-5 seconds prior to testing, and a calibrated pipette was used to transfer the sample volume specified in each manufacturer’s instructions for use.

### Ethical approved

This study was approved by the COVID-19 Scientific Research Committee of Ministry of Health (S.B. 2020-05-13T14_11_33) and by local Non-Interventional Research Ethics Committee of Kocaeli University (GOKAEK-2020/08.30 - 2020/144).

### Statistical analysis

The chi-square tests were calculated using a significance level of 95%. Chi-square scores were calculated using Excel. The heatmap is obtained by comparing chi-square scores for a range of Ct threshold combinations, where any patient with Ct value below the threshold is assumed positive for Bio-speedy and any patient with Ct value below the threshold for both of the genes (gene E and RdRp) is assumed positive for Diagnovital.

For Deming regressions, the data pairs with only positive Ct values were used for each experiment, except cumulatives. To obtain the cumulative Ct scores, an arbitrary copy number was obtained using 2^-Ct^ for gene E and RdRp obtained by Diagnovital, where the Ct is available. Otherwise, an arbitrary copy number of zero is used. The arbitrary copy numbers were added to obtain a cumulative copy number, and the cumulative Ct is obtained by ln (cumulative copy number)/ln(2). Calculations were done using Excel.

## RESULTS

Of all analyzed samples, 68 were positive, and 28 were negative by two PCR kits. The mean age of the study population was 44 (range;17 - 85), and 67 (64%) were male, including both genders showing signs and/or symptoms of COVID-19 infection. Demographic, laboratory, and medical findings of the study patients are shown in Table 2.

**Table 2.** Demographic, laboratory and medical findings of the study patients

| **Characteristic** | **Patient group** |
| --- | --- |
| **Patient, n** | 96 |
| **Gender, M/F, n (%)** | 67 (64)/32 (33) |
| **Age, years mean, range** | 44 (17-85) |
| **Sampling, region/city** | Marmara/Kocaeli |
| **Clinical status, n (%)** |  |
| **Symptomatic patients** | **62 (65)** |
| Cough | 15 (24) |
| Fever + cough + headache + sore throat | 9 (14) |
| Cough + respiratory distress | 7 (11) |
| Fever | 6 (10) |
| Sore throat + cough | 6 (10) |
| Fever + cough | 5 (8) |
| Cough + fatigue + headache | 3 (5) |
| Fever + fatigue + myalgia | 3 (5) |
| Fever + cough + malaise | 2 (3) |
| Malaise + sore throat + myalgia | 2 (3) |
| Cough + fatigue + sore throat | 2 (3) |
| Fever + cough + loss of taste | 1 (2) |
| Fever + abdominal pain + vomiting | 1 (2) |
| **Asymptomatic patients** | **34 (35)** |
| **Working status, n (%)** | **50 (52)** |
| Worker | 20 (40) |
| Self-employed | 8 (16) |
| Housewife | 6 (12) |
| Public personnel | 6 (12) |
| Retired | 5 (10) |
| Health employee | 4 (8) |
| Unemployed | 1 (2) |
| **Cormorbidity, n (%)** | **12 (13)** |
| Hypertension | 4 (33) |
| Diabetes mellitus | 3 (25) |
| Diabetes mellitus + hypertension | 2 (17) |
| Chronic obstructive lung diseases | 2 (17) |
| Cardiac diseases | 1 (8) |
| **Computerized tomography status, n (%)** |  |
| CT positive patient | 29 (30) |
| CT negative patient | 37 ((39) |
| CT not done | 30 (31) |

Abbreviations: M: male; F: female; CT: computerized tomography; PCR: polymerase chain reaction

Diagnovital and Bio-speedy kits were compared by RT-PCR amplification yield results. The initial quantity and yield quality of targets had been monitored by the exponential amplification phase of the RT-PCR reaction. There were different Cts for the same patient samples, and each kit indicated differently yield quality. Especially, Diagnovital showed in a constricted range and conveniently exponential amplification curves than Bio-speedy (Fig 1).

**Figure 1.**
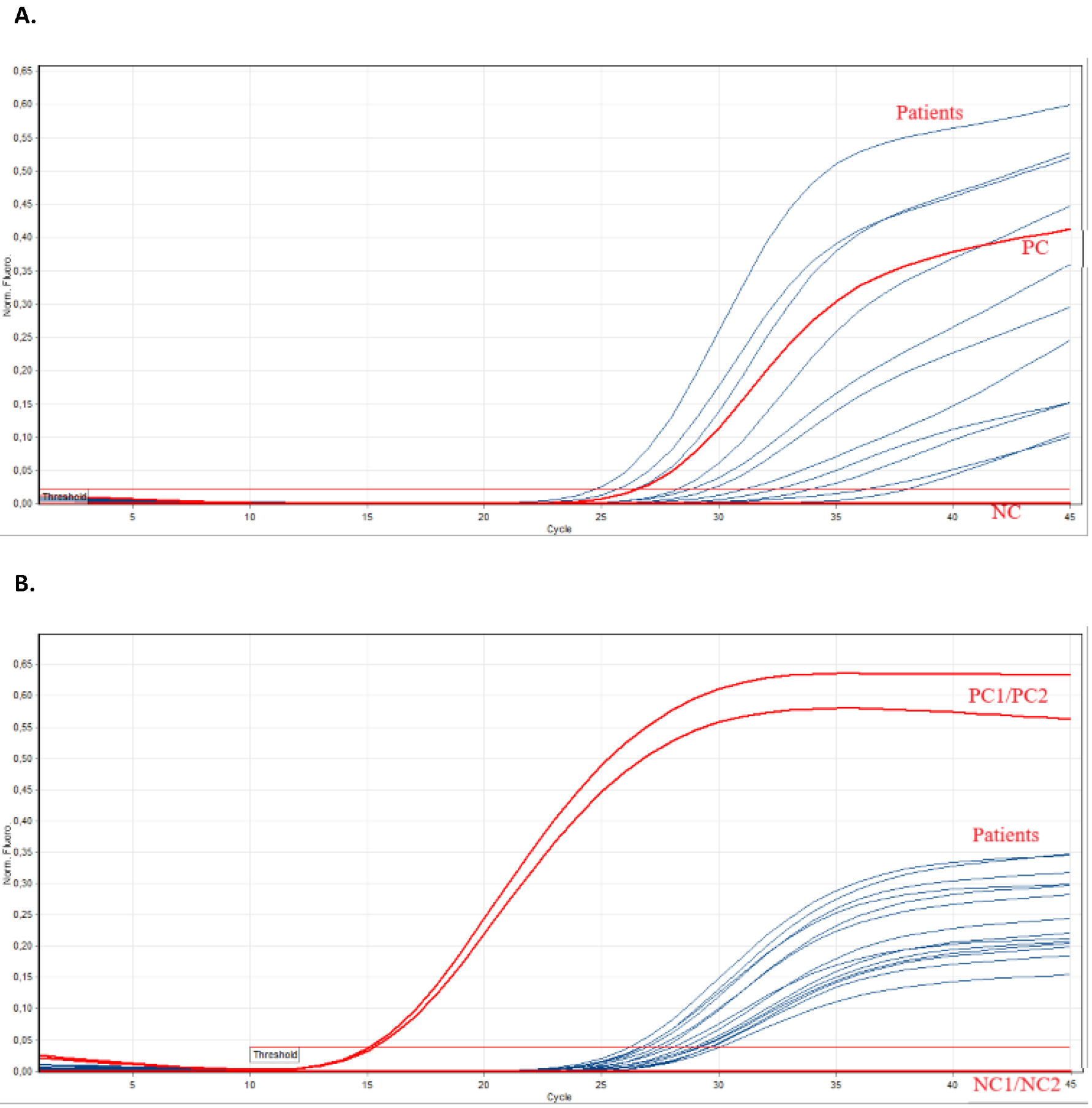
**A**. Revers transcriptase - polymerase chain reaction amplification yield results. BioSpeedy® SARS-CoV-2 PCR (RdRp) kit (upper) and, **B**. Diagnovital® SARS-CoV-2 PCR (RdRp) kit (lower) in Rotor-Gene Q Software 2.3.1. (Abbrevations: PC; positive control, NC; negative control).

The Deming regression of Ct values obtained for RdRp gene for the two separate kits and 95% confidence interval for the regression line shown in Fig 2. Diagnovital and Bio-speedy both use RdRp as a biomarker for their detection. The regression of Cts obtained for this biomarker showed correlation. It was important to note that since the isolation methods and possibly primers were different, we did not expect to observe a slope of 1. Indeed, Bio-speedy showed higher variation when it comes to Cts, yet the correlation was statistically significant (p<0.01) with a Pearson correlation of 0.73 (Fig 2).

**Figure 2.**
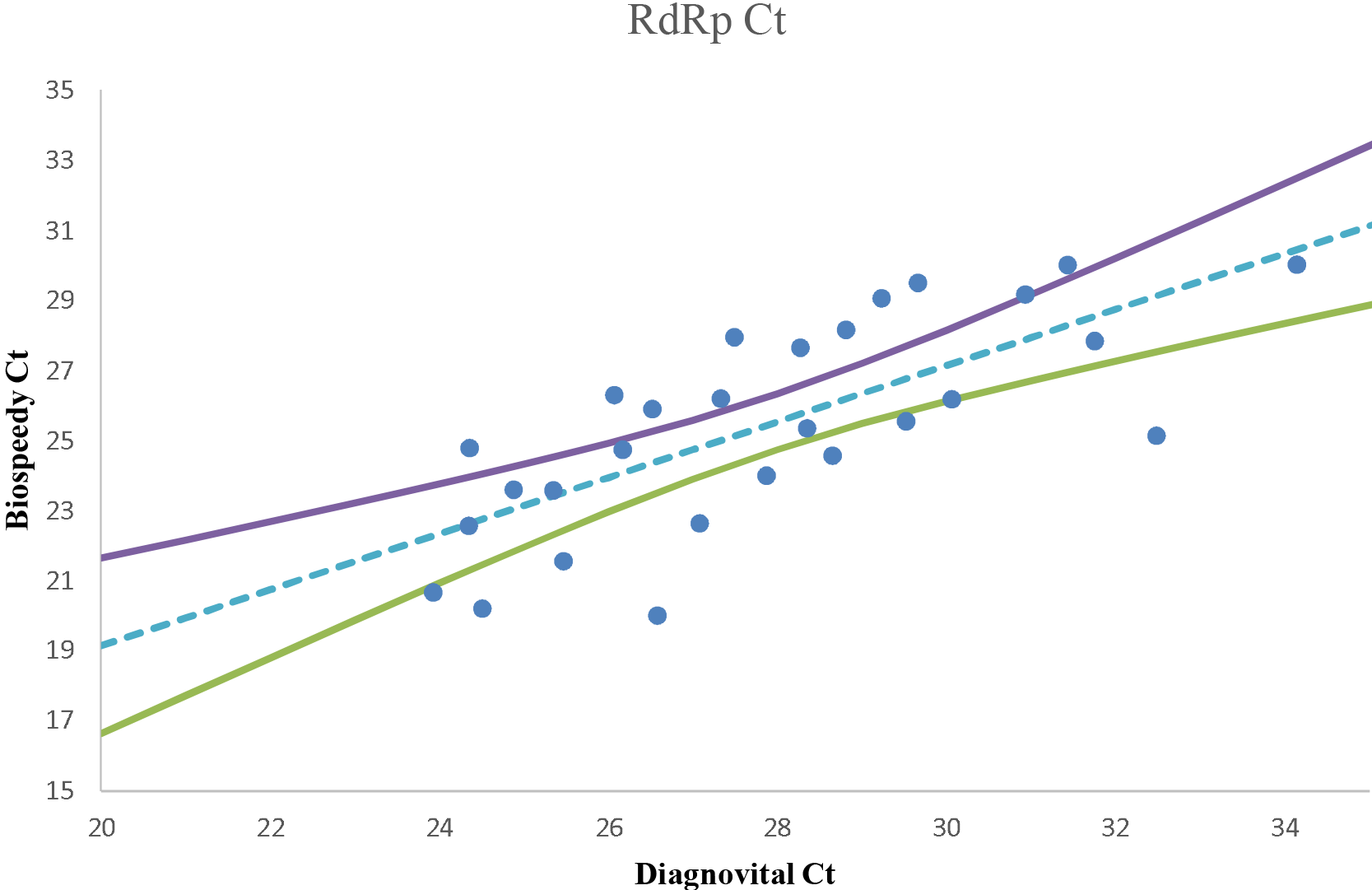
The Deming regression of Ct values obtained for RdRp gene for the two separate kits and 95% confidence interval for the regression line. Any data pair with any negative results are omitted.

The Deming regression of Ct values obtained for RdRp gene and gene E for Diagnovital and 95% confidence interval for the regression line shown in Fig 3. The two biomarkers used by Diagnovital, gene E, and RdRp gene were also showed some degree of correlation. This correlation was found statistically significant (p<0.01) with a Pearson correlation of 0.82 (Fig 3).

**Figure 3.**
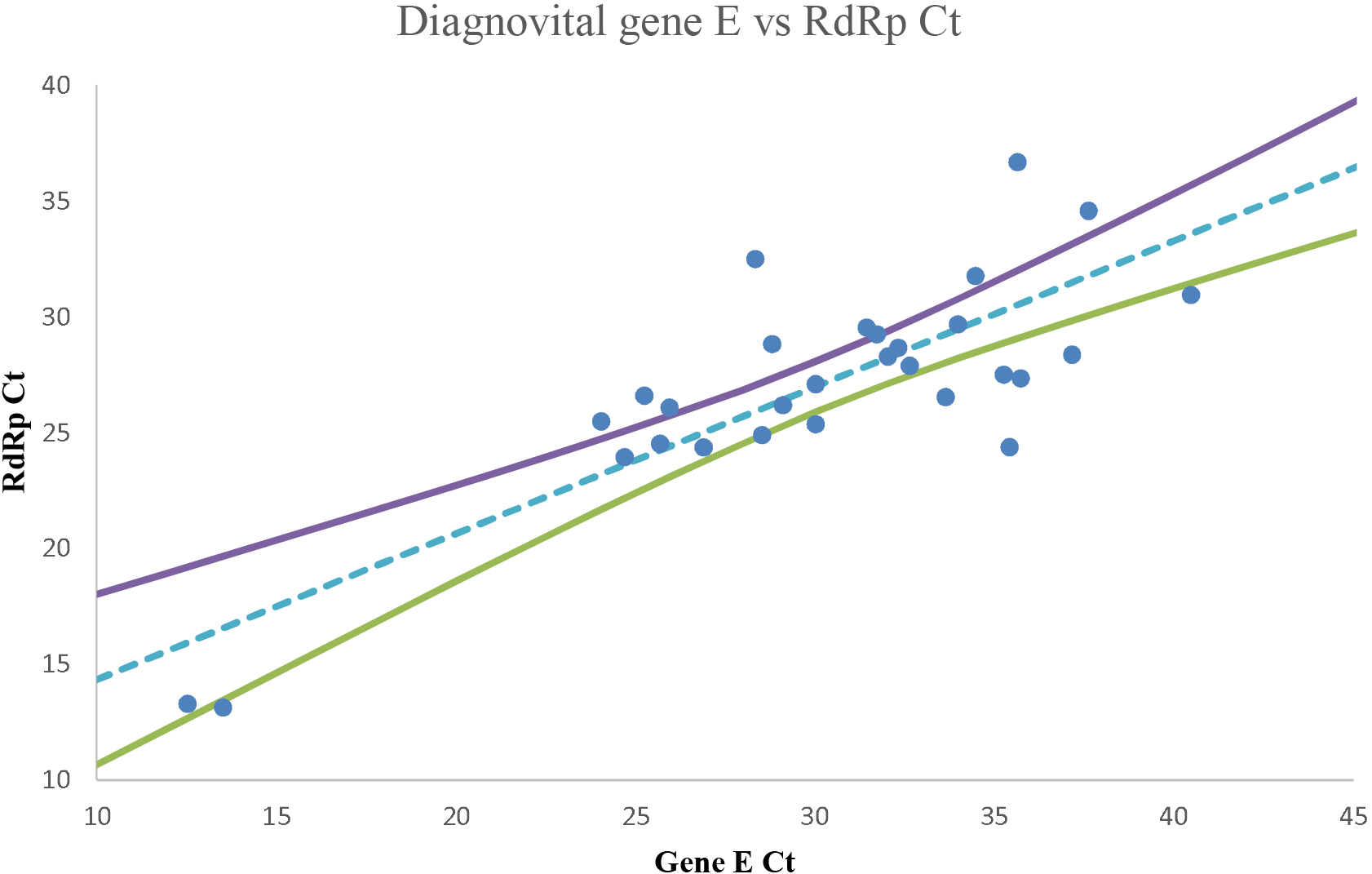
The Deming regression of Ct values obtained for RdRp gene and gene E for Diagnovital and 95% confidence interval for the regression line. Any data pair with any negative results are omitted.

Fig 4 shows **t**he Deming regression of the cumulative Ct values of Diagnovital and RdRp Ct values of Bio-speedy and 95% confidence interval for the regression line. The cumulative Ct values were calculated by the sum of arbitrary copy numbers of gene E and RdRp genes obtained through 2^-Ct^. The cumulative scores, when compared to Bio-speedy results, showed statistically significant (p<0.01) correlation with a Pearson coefficient of 0.83. This value being higher than 0.73 coefficient obtained through comparison of RdRps of the two kits only, showed that inclusion of a secondary biomarker by Diagnovital improved the correlation of different kits.

**Figure 4.**
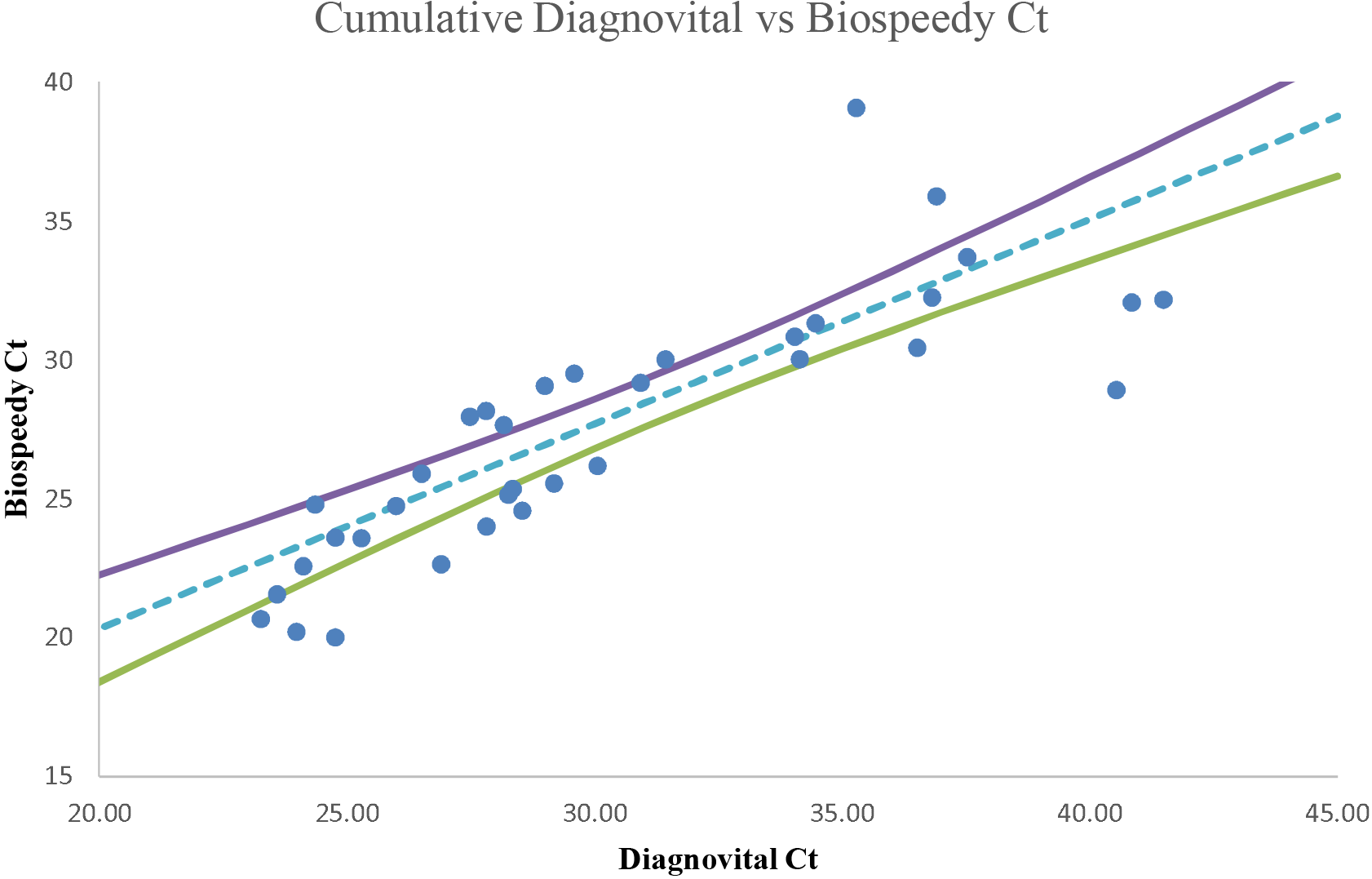
The Deming regression of the cumulative Ct values of Diagnovital and RdRp Ct values of Biospeedy and 95% confidence interval for the regression line. The cumulative Ct values were calculated by the sum of arbitrary copy numbers of gene E and RdRp genes obtained through 2^-Ct^. The arbitrary copy numbers for negatives were assumed as zero.

In Fig 5 the analyzed heat map for chi-square scores obtained for a range of thresholds for Bio-speedy and Diagnovital kits are shown. Black and white squares indicateD chi-square score of 1 and 0, respectively. According to chi-square heat map, while the two kits showed high independence for manufacturers choice of Ct, there was a range of Ct combinations for the two where the two kits showed a high correlation. When the Ct of 40 for Bio-speedy and 35 for Diagnovital was applied as instructed by the manufacturers, the chi-square score of 0.18 was obtained, indicating a high independence between the kits for manufacturer defined Cts. On the other hand, the maximum dependence for the two kits could be established if the Ct were modified to any of the 38 and 32, 38 and 34, 37 and 35, 34 and 37 combinations for Bio-speedy and Diagnovital, respectively. While dependence could increase at even higher Ct values, it is important to note that high dependence is not an indication of high sensitivity or specificity.

**Figure 5.**
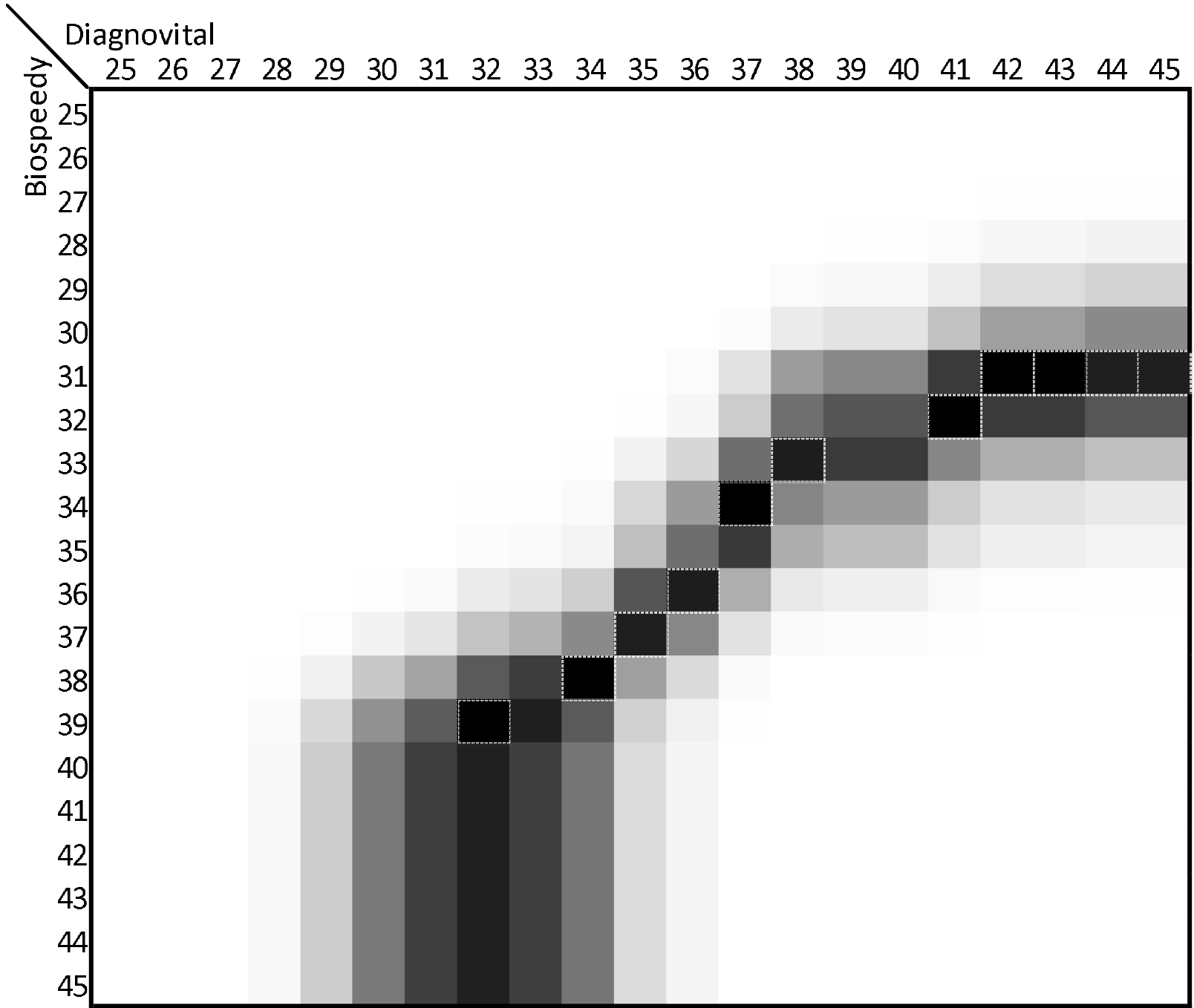
Heat map for chi-square scores obtained for a range of thresholds for Biospeedy and Diagnovital kits. Black and white squares indicate chi-square score of 1 and 0, respectively. The black squares with white borders indicate Ct threshold combinations where chi square scores are above 0.88.

## DISCUSSION

On January 26, the first batch of four registration certificates of Novel Coronavirus PCR kit was issued by the State Food and Drug Administration for the emergency, without a series of clinical trials (https://www.fda.gov/news-events/press-announcements/coronavirus-covid-19-update-fdaissues-new-policy-help-expedite-vailability-diagnostics). Then, new PCR kits began to be produced by different laboratories and institutes. Many studies on the obtained kits have also been carried out and are still underway (22). Nowadays, Turkey is among the countries producing SARS-CoV-2 diagnostic PCR kits. In this study, we investigated the compatibility between the two different SARS-CoV-2 PCR kits, which included different gene targets that produced in Turkey during the COVID-19 pandemic. We used two separate SARS-CoV-2 RT-PCR kits similar to each other, including the procedure, analytic sensitivity, storage of the kits, studied samples, and other features shown in Table 1. The differences were the addition of E gene analysis in addition to RdRp gene analysis in the Diagnovital PCR kit and isolation methods. In the absence of international quality controls, an unknown sample may more accurately diagnose with a high cumulative correlation of kits. According to two PCR kits, there were 68 positive and 28 negative results as cumulative in our study.

The mean age of our patient population was 44, and 64% of them were male. There are many articles reporting gender differences in terms of frequency and severity of the disease (23,24,25). The higher prevalence is in males because ACE2 expression is more dominant in men, and smoking is more common in men than in women (26). The most common symptoms on admission were cough (24%), 10% fever, and there was more than one symptom association. Within 68 positive patients, there were 15 patients (22%) without symptoms (data not shown). Studies are showing variable results about the transmission rates of asymptomatic individuals (27,28). The samples came from the city of Kocaeli, one among the cosmopolitan cities of the country, where the industry is developed, and the working class is quite high. The COVID-19 epidemiology varies between regions. Among the possible reasons for this variation are the demographic and socio-cultural structure, transportation. The city of Kocaeli is also among the cities where the country’s export and import are made the most. That is why the number of workers was considerably high (40%) in our study population. Comorbidities were present in 13% of the patients. The comorbidities with the highest frequency were as follows: coronary artery disease with hypertension (33%), diabetes (25%), chronic obstructive lung diseases (17%). Similar results for comorbid diseases were obtained in a study from China. (29). In 5 of 28 PCR (18%) negative patients, CT findings were consistent with COVID-19, and all five patients had symptoms (data not shown).

It was interesting that different Cts for the same patient samples were seen in each kit. Starting quantities were the same, but they had different yield quality. Especially, Diagnovital PCR kit showed in a constricted range and conveniently exponential amplification curves than Bio-speedy PCR kit (Fig 1). This case suggested that two different RNA target gene assays were more appropriate as suggested by WHO in the diagnosis of COVID-19 disease (15). In general, it is difficult to understand the analytical sensitivity differences of different molecular tests due to natural variations in sample processing and reference materials used for validation in different laboratories. However, the study comparing three different molecule tests by Uhteg K et al. showed that similar results were found; The PCR kit with two different genes of the SARS-CoV-2 had a higher yield than the other two kits performing one gene analysis (30).

Our data suggested that all two PCR methods yielded comparable results (a Pearson coefficient of 0.83) for both negative and positive clinical specimens (Fig 4). However, we did find a notable difference comparison of RdRps of the two kits had a correlation of 0.73 (Fig 2). It was important to note that since the isolation methods and possibly primers were different, we did not expect to observe a slope of 1. Indeed, Bio-speedy showed higher variation when it comes to Cts, yet the correlation was statistically significant (p<0.01). This case indicated that while the two kits reported similar results for the RdRp gene, there was a considerable difference as well. The addition of a secondary biomarker for Diagnovital improved the correlation of different kits and increased the correlation by 0.10 (Fig 4). While Bio-speedy did not alone represent the true values, the increased correlation was an indication of improved diagnostic power. On the other hand, as shown in Figure 3, both biomarkers were somewhat correlated with the 95% confidence interval for the Deming regression line of the Ct values obtained for the Diagnovital for the RdRp gene and the E gene. This result indicated that the two biomarkers reported relatively close results adding to the cumulative detection power.

Choosing a threshold level is essential to finalize the diagnosis and classify the patient. Most diagnostic tools refer to any Ct value above a specific threshold, too low for detection and refrain from a conclusive classification. In the case of a pandemic event, the doctors, however, do not have the luxury or resources. Unfortunately, without true positives or true negatives, we do not have the means to perform ROC analyses in order to determine the right thresholds to reach an optimum diagnostic power. In spite, we had the means to compare the two diagnostic tools and how they were correlated with each other. Using Deming, while it was possible to correlate the Cts; however, the thresholds for diagnosis could not be correlated as a result of diagnosis was categorical rather than continuous data, as in Cts. Instead, we had the means of correlation of categorical data using the chi-square score as a specific threshold combination. However, depending on the choice of thresholds, this score was prone to change. During a grid search, we scanned a range of thresholds for both of the kits, producing a heatmap of chi-square scores (Fig 5). Important to note that the heatmap did not provide information on accuracy, but rather, correlation of the kits. According to the chi-squares heatmap, the range of high correlation thresholds is along the diagonal of Ct ranges. Surprisingly the correlation dramatically dropped outside of this diagonal, including the threshold combination determined by the kit manufacturers (35 for Diagnovital and 40 for Bio-Speddy). This case indicated that while the correlation for Cts exists, the choice of threshold might strongly influence the diagnosis. To obtain a higher accuracy, a combinatory study was advisable to determine optimum thresholds using multiple kits for even a few hundred cases.

The RT-PCR tests today are so new that it is unclear how reliable they are. Local laboratories are doing their best to validate tests on their own. They are also aimed at scanning more masses by reaching more tests and thus preventing the spread of the disease by detecting positives. In our country Scientific and Technological Research Council of Turkey had also accepted all projects about PCR kits. This was conducted because the country needed these kits to help with quality assurance internally. A viewpoint published in JAMA earlier this month synthesized known data on the accuracy of different tests at different points in the disease process (31). Some kits have three target genes, use one target gene for the screening test and two other target genes for verification test. The confirmatory test result is considered positive only when both confirmatory genes are detected. If a gene is not detected, the result cannot be interpreted positively (21).

In conclusion, it should be mentioned that the international external quality controls for the diagnosis of SARS-CoV-2 by PCR techniques used have not been established yet. Our results suggest, in an unknown sample on the light of clinic symptoms of COVID-19 using together different PCR kits that target different genes during the pandemic situation may provide a more accurate diagnosis.

## Data Availability

Data were collected by Sila Akhan and Murat Sayan.

## Acknowledgement

We are thankful to Tarik Keçeli for English editing.

